# Mid-Life Plasmalogens and Other Metabolites with Anti-Inflammatory Properties are Inversely Associated with Long term Cardiovascular Disease Events: Heart SCORE Study

**DOI:** 10.1101/2023.03.02.23286731

**Authors:** Anum Saeed, Chris McKennan, Jiaxuan Duan, Kevin E. Kip, David Finegold, Michael Vu, Justin Swanson, Oscar Lopez, Ann Cohen, Mark Mapstone, Steven E. Reis

**Affiliations:** University of Pittsburgh School of Medicine, Pittsburgh, PA; Heart and Vascular Institute, UPMC, Pittsburgh, PA; University of Pittsburgh, Department of Statistics, Pittsburgh, PA; Clinical Analytics, UPMC Health Services Division, Pittsburgh, PA; University of Pittsburgh School of Public Health, Pittsburgh, PA; University of California, Irvine, Department of Neurology, Irvine, CA; University of South Florida, Tampa, FL; Cognitive and Behavioral and Neurology Division, UPMC, Pittsburgh, PA

## Abstract

**Background:** Preclinical data have shown that low levels of plasmalogens and other metabolites with anti-inflammatory properties may impact metabolic disease processes. However, the association between mid-life levels of such metabolites and late-life atherosclerotic cardiovascular disease (ASCVD) is not known.

**Methods:** We characterized the midlife plasma metabolomic profile (1,228 metabolites) of 1,852 participants (age 58.1±7.5 years, 69.6% female, 43.6% self-identified as Black) enrolled in the Heart Strategies Concentrating on Risk Evaluation (Heart SCORE) study. Participants were followed for ∼16 years for incident ASCVD events (nonfatal MI, acute ischemic syndrome, coronary revascularization and ASCVD mortality). We used regression model to assess associations of metabolites with ASCVD events. We assessed the impact of genetic variants using whole-exome sequencing with single-variant analysis for common variants and gene-based burden tests for rare variants. We used unbiased and candidate gene approaches to explore genetic associations with metabolites found to be associated with ASCVD events.

**Results:** A total of twelve metabolites were independently associated with incident ASCVD in fully adjusted models over a median of 12.1 years. A subset of plasmalogens showed an independent inverse association with incident ASCVD events [1-(1-enyl-palmitoyl)-2-arachidonoyl-GPC (OR, 0.54; 95% CI, 0.40-0.74); 1-(1-enyl-palmitoyl)-2-arachidonoyl-GPE (OR, 0.57; 95% CI, 0.42-0.78), 1-methylnicotinamide1-(1-enyl-stearoyl)-2-arachidonoyl-GPE (OR, 0.76; 95% CI, 0.65-0.89)]. Metabolome-wide genetic analysis revealed that two of these plasmalogen metabolites were strongly influenced by polymorphisms of the rs174535, an eQTL for FADS1 and FADS2 genotype. Two amino acid metabolites (2-oxoarginine [OR, 0.42; 95% CI, 0.25-0.69], alpha-ketobutyrate [OR, 0.62; 95% CI, 0.49-0.80]) and a bilirubin degradation product (C_16_H_18_N_2_O_5_ [OR, 0.50; 95% CI, 0.38-0.66) were inversely associated with ASCVD events.

**Conclusions:** Higher mid-life levels of three plasmalogens, two amino acid metabolites, and a bilirubin degradation product, all of which have anti-inflammatory properties, are associated with lower risk of late-life ASCVD events. Further research is needed to determine whether these metabolites play a causal role in ASCVD and may be a target for future therapies.

## INTRODUCTION

Atherosclerotic cardiovascular disease (ASCVD) is a leading cause of mortality and morbidity worldwide (1). The process of atherosclerosis begins decades before clinical manifestations of ASCVD events (2,3). Therefore, identification of pathophysiologic and protective markers of atherosclerosis in mid-life is essential in primordial and primary prevention of ASCVD events in late-life (4,5). These markers can also provide insights into the development of novel targets for developing preventative therapies.

It is well known that inflammation-induced oxidative stress, dysregulated lipid metabolism and elevated remnant cholesterol contribute to the residual risk of cardiovascular disease (CVD). This residual risk is beyond what is captured by traditional lipid profiles (6) and clinically available risk assessment tools. The use of metabolomic analyses for comprehensive profiling of molecular markers in the lipid, amino acid and other pathways in systemic disease processes (7-9) provides an opportunity to identify the role of these metabolites in the development of ASCVD.

Preclinical data indicate that low levels of plasmalogens, a class of membrane glycerophospholipids, modulate cardiometabolic changes (10) which could lead to atherosclerosis. However, population level associations of mid-life plasmalogens and other metabolites with anti-inflammatory properties with incident ASCVD events are lacking. Here, we assessed midlife metabolomic architecture with risk of incident ASCVD events over a mean of 10.4 years of follow-up in the Heart Strategies Concentrating on Risk Evaluation (Heart SCORE) study. We then investigated genetic level associations with metabolites found to be related with incident ASCVD events.

## METHODS

### Study population

The Heart Strategies Concentrating on Risk Evaluation (Heart SCORE) study began in 2003 as a community-based participatory research study of race-related disparities in ASCVD conducted in Allegheny County, Pennsylvania. A detailed description of the Heart SCORE study design has been published elsewhere (11,12). The study enrolled 2,000 participants age 45 to 75 years. Those with a life expectancy <5 years or an inability to undergo annual follow-up visits were excluded.

### Assessment of Covariates

All participants provided written informed consent approved by the University of Pittsburgh Institutional Review Board. Baseline evaluation included assessment of demographics, psychosocial characteristics, anthropometric measurements, exercise and dietary habits, traditional ASCVD risk factors, and sleep disturbances. Diabetes mellitus was defined as a fasting serum glucose level ≥126 mg/dL or nonfasting serum glucose level ≥200 mg/dL, a self-reported physician diagnosis of diabetes, or use of hypoglycemic medication. Participants were classified as “never,” “former,” or “current” smokers from self-reported information. Blood pressure (BP) was measured by trained technicians following a standard protocol. Fasting venous lipid profile was measured using standard enzymatic methods (13) at the University of Pittsburgh Medical Center clinical laboratory. Interleukin (IL-6) levels were measured using commercially available ELISA kits (Quantikine high-sensitivity [hs] human kits, R&D Systems, Minneapolis, Minn). High-sensitivity C-reactive protein (hs-CRP) was measured by a high-sensitivity method on a Hitachi 911 analyzer (Roche Diagnostics, Basel, Switzerland) using reagents from Denka Seiken (Niigata, Japan). Baseline plasma samples were stored for future analyses. The present analyses were confined to 1,852 participants who self-reported race as either Black or White.

### Coronary artery calcium measurement

Electron beam computed tomography image acquisition was obtained using an Imatron C150 scanner (GE Imatron Inc., South San Francisco, California) to scan 3-mm of the heart during a single inspiratory breath-hold. Calcium scores were calculated by the Agatston method using a densitometric program (14). An experienced reader blinded to subject identities interpreted scans (15).

### Metabolite profiling

Metabolomic profiling was completed on plasma from samples obtained in the fasting state at the baseline visit and stored at −80°C. Duplicate technical replicates were done for 99 participants for validation purposes, where one replicate was chosen at random to be used in all downstream analyses. Equal volumes of all samples were extracted and run across the Metabolon Inc. (Durham, NC) Precision Metabolomics discovery platform (16,17). Samples were extracted and split into equal parts for analysis on four LC/MS/MS platforms. Proprietary software was used to match ions to an in-house library of standards for metabolite identification and for metabolite quantitation by peak area integration, where a metabolite’s abundance was defined as its peak area. Metabolites with >10% missing data were discarded to avoid missing data-related biases.

### Genetic Profiling

Blood samples for genotyping were collected in 10mmol/L EDTA. DNA was isolated following standard protocols. A customized Illumina CARe iSelect (IBC) cardiovascular array developed for cardiometabolic phenotypes (18) was used for genotyping. The IBC array is a 50K SNP chip that assays polymorphisms in 2100 genes.

### Incident Cardiovascular Disease Events

Ongoing annual evaluations include measurement of risk factors, tabulation and adjudication of adverse events, and periodic assessments of subclinical atherosclerosis. The primary outcome was a composite of ASCVD mortality and nonfatal ASCVD events, defined as nonfatal myocardial infarction, acute ischemic syndrome, coronary revascularization (percutaneous coronary intervention or coronary artery bypass graft). All events were tabulated by annual telephonic or in-person follow up, confirmed by hospital record review, and adjudicated by staff cardiologists. Mortality was ascertained and classified as ASCVD death by reviewing death certificates and hospital records. Outcomes were analyzed until December 31, 2020 with a median of 12.1 (9.2, 12.3) years follow up period.

### Statistical analyses

#### Baseline Comparisons

Baseline characteristics between Black and White participants are compared using the Welch Two Sample t-test and Pearson’s Chi-squared tests for categorical and continuous variables, respectively.

#### Identification of Cardiovascular Disease Metabolite Associations

Latent factors that might confound the relationship between metabolite levels and ASCVD events were estimated using the method described by McKennan, et al. (19). For each metabolite, we used logistic regression to regress ASCVD (yes/no) onto that metabolite’s log-abundance while controlling for the baseline covariates in two models. First, we used the “*Basic model*,” including the variables of the Pooled Cohorts Equations (PCE)(age, sex, race, hypertension medication use, systolic BP, smoking status, diabetes mellitus, high density lipoprotein (HDL) cholesterol, total cholesterol) (20,21). We then controlled for inflammatory markers by using a second “*Complete model*,” which included the covariates in the Basic model and log-concentrations of hs-CRP and IL-6. Both models were adjusted for latent factors. The Benjamini-Hochberg procedure (22) was used to control the false discovery rate (FDR) at 10%.

#### Metabolite Genome-Wide Association Study (GWAS)

DNA samples from 1,132 Heart SCORE participants with metabolite, baseline covariate information was used to perform GWAS analyses. Quality control (QC) was performed by removing SNPs with >10% missing data, with minor allele frequency <1%, or in Hardy–Weinberg disequilibrium (p-value≤10^−5^) in either self-identified whites or blacks, which resulted in 43,465 candidate SNPs. The method of Zhao, et al. (23) was used to perform two independent metabolomic GWAS’s, which estimate indirect and direct genetic effects on metabolite levels where indirect effects are mediated through latent factors and the direct effect analysis is equivalent to standard latent factor-corrected quantitative trait loci analyses (24). Adjustments were made for baseline covariates and percent African ancestry in each GWAS. Metabolite-SNP or Plasmalogen-SNP pairs with p-values ≤0.05/(#Metabolites^*^#SNPs) or ≤0.05/(#Plasmalogens^*^#SNPs) from either GWAS were considered metabolome-wide or Plasmalogen-wide significant, respectively.

## RESULTS

The 1,852 participants were 59.1±7.5 years of age at study entry (Table 1). Women comprised 65.8% of the cohort; 43.6% of participants self-identified as Black. Black participants had a higher prevalence of high Framingham risk (25.1 vs. 13.7%), were more likely to be smokers (14.4 vs 8%), and had higher prevalence of hypertension (55.9 vs 38.3%) and diabetes (15.8 vs 5.3%) as compared to Whites. White participants had nominally higher total cholesterol levels (216 vs 209 mg/dL). Of the subset 812 participants who underwent coronary artery calcium scans at baseline, more Black participants had an Agatston CAC score >0 (32.7 vs 30.2%; p<0.01). However, there were fewer Black participants with a CAC>100 (9.8% vs 14.8%; p<0.01).

**Table 1:** Baseline Characteristics Stratified by Self-Reported Race.

|  | **Overall**<br>**(N=1852)** | **Black**<br>**(N=808)** | **White**<br>**(N=1044)** | **p-value** |
| --- | --- | --- | --- | --- |
| **Age, y** | 59.1 (7.5) | 58.1 (7.5) | 59.8 (7.4) | <0.001 |
| **Female** | 1218 (65.8%) | 562 (69.6%) | 656 (62.8%) | 0.003 |
| **BMI, Mean**<br>**(SD)** | 30.1 (6.2) | 32.0 (6.4) | 28.6 (5.5) | <0.001 |
| **Hypertension** | 852 (46.0%) | 452 (55.9%) | 400 (38.3%) | <0.001 |
| **Systolic BP**<br>**(mmHg)** | 137 (19.8) | 141 (20.1) | 133 (18.8) | <0.001 |
| **Current**<br>**Smoking** | 200 (10.8%) | 116 (14.4%) | 84 (8.0%) | <0.001 |
| **Diabetes**<br>**Mellitus** | 183 (9.9%) | 128 (15.8%) | 55 (5.3%) | <0.001 |
| **Total**<br>**Cholesterol,**<br>**mg/dL** | 213 (42.7) | 209 (44.4) | 216 (41.1) | <0.001 |
| **HDL-C,**<br>**mg/dL** | 57.6 (14.9) | 58.1 (14.4) | 57.2 (15.3) | 0.2 |
| **Hs-CRP**<br>**mg/dL** | 0.363 (1.2) | 0.63 (1.2) | 0.16 (1.2) | <0.001 |
| **IL-6 pg/mL** | 0.525 (0.75) | 0.78 (0.68) | 0.36 (0.75) | <0.001 |

Data are displayed as mean±SD for continuous variables and n (%) for categorical variables.
Abbreviations: SD, standard deviation; BMI, body mass index; BP, blood pressure; HDL-C, high-density lipoprotein cholesterol; HS-CRP, high sensitivity C reactive protein, IL-6, Interleukin-6.

**Table 2:** Metabolites Associated with CVD events and Corresponding Genetic Polymorphisms.

| Cardiovascular Disease Event Risk |  |  |  |
| --- | --- | --- | --- |
| Metabolite | Odds Ratio<br>95% CI | FDR adjusted p-value | Gene Polymorphism<br>Influence |
| 1-(1-enyl-palmitoyl)-2-arachidonoyl-GPC<br>(P-16:0/20:4) | 0.54<br>95% CI, 0.40-0.74;<br>p<0.001 | 0.04 | FADS1/FADS<br>(*GWAS in HeartSCORE*) |
| 1-(1-enyl-stearoyl)-2-arachidonoyl-GPE<br>(P-18:0/20:4) | 0.57<br>95% CI, 0.52-0.78;<br>p<0.001 | 0.08 | FADS1/FADS2<br>(*GWAS in HeartSCORE*) |
| C16H18N2O5(2) | 0.50<br>95% CI, 0.38-0.66<br>p<0.0001 | 0.0005 | UGT1A<br>(*GWAS in HeartSCORE*) |

The composite endpoint of incident ASCVD events occurred in 2.9% of participants followed for median of 12.1 (9.2, 12.3) years. This included ASCVD mortality (2.9%) and incident MI (2.6%). Cardiac mortality was noted to be higher in Black compared to White participants (3.8% vs. 2.2%, respectively; p-value=0.04).

### Distribution of Metabolites in Heart SCORE

A condensed Heat map of the Heart SCORE cohort metabolite distribution with both gender and racial comparisons is included as <u>Supplemental Figure i.</u> In both Blacks and Whites, males had higher levels of amino acid-associated metabolites and lower levels of many lipid species, with the exception of acyl carnitines and steroids. For example, males had higher levels of pregnenolone, androgens, and corticosteroids while females had higher levels of progestins. Notably, while medium-chain (MCFA), long-chain (LCFA), and polyunsaturated (PUFA) free fatty acids levels were lower in males, levels of acylcarnitines were higher. Black participants were noted to have lower levels of MCFA, LCFA, and PUFAs compared to Whites. Plasmalogen levels were nominally higher in Black participants at baseline compared to White participants.

### Identification of Metabolites Associated with Incident ASCVD Events

We performed regression analyses of the full panel of 1,228 metabolites using the Basic and Complete models. Figure 1A is a Manhattan-like plot under the complete model, where 12 metabolites were significantly associated with incident ASCVD events at a 10% FDR (Figure 1B). Of these, 6 metabolites were from the lipid pathway (three Plasmalogens species, two Phosphatidylinositol and Phosphatidylethanolamine species, one long chain fatty acid), two were from the amino acid pathway (alpha ketobutyrate, 2-oxarginine), one was a bilirubin degradation product (C16H18N2O5), and three were unnamed.

**Figure 1:**
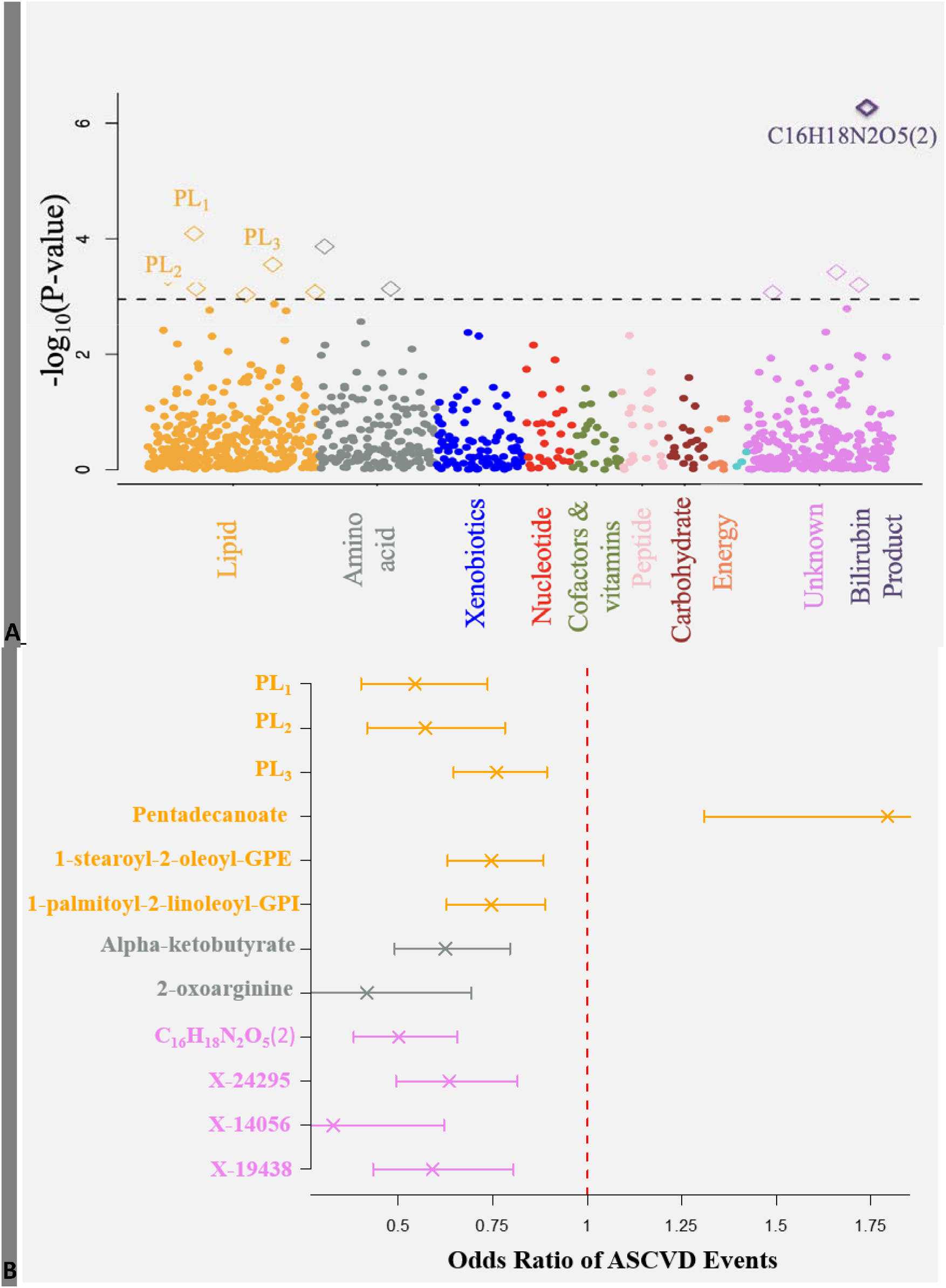
Plasma Metabolites Pathways and Risk for Incident Atherosclerotic Cardiovascular Dis (ASCVD) Events. **Figure 1A:** Manhattan Plot for pathway analysis for metabolites’ associations with risk for ASCVD events, **Figure 1B:** Odds Ratios for incident ASCVD events (95% CI) per SD increment for midlife metabolites. Abbreviations: PL, Plasmalogens; PL1, 1-(1-enyl-palmitoyl)-2-arachidonoyl-GPC (P-16:0/20:4); PL2, 1-(1-enyl-palmitoyl)-2-arachidonoyl-GPE (P-16:0/20:4); PL3, 1-(1-enyl-stearoyl)-2-arachidonoyl-GPE (P-18:0/20:4)

Three Plasmalogens, a subclass of lipids, were found to be significantly inversely associated with incident ASCVD events (Fisher’s exact test p<0.001). These included: 1-(1-enyl-palmitoyl)-2-arachidonoyl-glycerophosphatidylcholine [GPC] (P-16:0/20:4) [OR, 0.54;95%CI, 0.40-0.74; p<0.001); 1-(1-enyl-palmitoyl)-2-arachidonoyl-glycerophosphatidylethanolamine [GPE] (P-16:0/20:4) [OR, 0.57; 95% CI, 0.42-0.78; p<0.001); and 1-methylnicotinamide1-(1-enyl-stearoyl)-2-arachidonoyl-GPE (P-18:0/20:4) [OR, 0.76; 95% CI, 0.65-0.89); p<0.001). Plasmalogens were significantly positively correlated with HDL cholesterol levels (all p-values<0.001).

Two metabolites in the amino acids pathway, 2-oxoarginine [OR, 0.42; 95% CI, 0.25-0.69; p<0.001] (an arginine metabolite) and alpha-ketobutyrate [OR, 0.62; 95% CI, 0.49-0.80; p<0.001] (a metabolite in the methionine and cysteine sub pathway), and a bilirubin degradation product (25), C16H18N2O5 [OR, 0.50; 95% CI, 0.38-0.66; p<0.001] showed a significant inverse association with ASCVD events. Two unidentified metabolites were similarly noted to have significantly reduced risk of ASCVD events.

### Metabolite Genome-Wide Association Study

We identified 641 metabolome-wide significant metabolite-SNP pairs, comprising 120 metabolites from 42 pathways and 193 SNPs from 74 genes. Notably, the abundances of two out of the three significant ASCVD-related Plasmalogens were associated with the genotype at rs174535 in FADS1/2 (Figure 3a). This genetic effect was less prominent in those with greater African ancestry (Figure 3b). This SNP is in near perfect linkage equilibrium with both rs174547 and rs174546 (r2=0.98 in Europeans), whose genotypes have been shown to modify aortic stenosis (25) and coronary artery disease risk (26), respectively. We also found the ASCVD-related metabolite C_16_H_18_N_2_O_5_ to be strongly associated with SNPs on UGT1A (Figure 3a), a regulator of bilirubin metabolism, which is consistent with C_16_H_18_N_2_O_5_ being a bilirubin degradation product.

## DISCUSSION

We identified that mid-life levels of several plasma anti-inflammatory and anti-oxidant metabolites are inversely associated with incident ASCVD events in late-life. Specifically, our data indicate that three *arachidonoyl* plasmalogens from the lipid sub pathway (1-(1-enyl-palmitoyl)-2-arachidonoyl-GPC, 1-(1-enyl-palmitoyl)-2-arachidonoyl-GPE and 1-methylnicotinamide1-(1-enyl-stearoyl)-2-arachidonoyl-GPE) are inversely associated with ASCVD risk over a median of 12.1 years of follow-up, independent of race, traditional ASCVD risk factors and inflammatory markers. Metabolome-wide genetic analysis revealed that two of these Plasmalogens are strongly influenced by polymorphisms on the FADS1/FADS2 gene, which are known to be associated with coronary artery disease and cardiometabolic health (26). Furthermore, two amino acid metabolites (2-oxoarginine and alpha-ketobutyrate) and a bilirubin degradation product (C_16_H_18_N_2_O_5_) were inversely associated with ASCVD events.

To our knowledge, this is the first report to show an inverse association of mid-life levels of bioactive plasmalogens with future ASCVD events. Our findings extend those of Meike et al. who, in a cross-sectional study of 220 patients, showed that phosphatidylethanolamine (PE) Plasmalogen species were inversely associated with unstable coronary artery disease but not with stable coronary disease (27). Plasmalogens are naturally occurring glycerophospholipids that are primarily present as phosphatidylcholine (PC) and phosphatidylethanolamine (PE) species (28). Previous studies have demonstrated that lower Plasmalogen levels are associated with inflammation-mediated aging diseases, such as Alzheimer’s disease (29). Therefore, we postulate that our findings of a cardioprotective role of plasmalogens may be related to their ability to prevent lipid oxidation (30), which modulates inflammation (31,32) and the initiation and progression of atherosclerosis (33) (28,34).

Recent data have shown the utility of supplementation of compounds with impact on oxidative stressors via multiple pathways could confer a risk reduction of ASCVD events (35). PE plasmalogens are known to prevent oxidation of cholesterol in phospholipid bilayers (36) and are more abundantly present in the cardiovascular system. plasmalogens’ anti-oxidant potential (10) may be due to the enhanced electron density of the vinyl ether bond at the sn-1 position that make it susceptible to cleavage by reactive oxygen species (ROS). The vinyl ether bond is preferably oxidized, while protecting the polyunsaturated fatty acids present in the sn-2 oxidation position (28,37,38). In a preclinical evaluation of cultured human pulmonary arterial endothelial cells, increasing the cellular Plasmalogen levels resulted in protection of cells during hypoxia and other ROS-mediated stress. The oxidative products of plasmalogens are unable to further propagate lipid peroxidation and may terminate the lipid oxidation process (39).

The biologic plausibility of our findings is supported by the results of our metabolome-wide genetic association analysis. Our results indicate that two of the three plasmalogen metabolites inversely associated with future ASCVD events were strongly influenced by the genotype at *rs174535* located on the *FADS1/FADS2* genes. While, located in the gene body of MYRF, rs174535 is only an expression quantitative trait loci (eQTL) for FADS1 and FADS2, where the latter lies 40k base pairs upstream from rs174535. While we did not find statistically significant associations between the SNPs on or near FADS1/FADS2 and ASCVD events, we observed that estimates were in the expected direction, with more copies of *rs174535*’s G allele rendering a greater risk of ASCVD. Although this is the first time such an association has been shown, FADS1/FADS2-related SNPs have been linked with lipid level regulation (40,41) (42). In addition, Yang et.al, showed the T allele at either *rs174546* or *rs174601*, which are in strong linkage disequilibrium with *rs174535* (r^2^>0.8 in Europeans), increases one’s risk for coronary artery disease and ischemic stroke. Other studies have shown that the *rs174546* T allele is located within the microRNA target site and associated with lower FADS1 mRNA levels in human liver (43).

We found that participants with African ancestry had a higher levels of plasmalogens (supplemental Table i) and the FADS genetic effect was smaller than that observed in those with European ancestry (Figure 2). As shown in Figure 2, *rs174535*, the SNP most associated with 1-(1-enyl-palmitoyl)-2-arachidonoyl-GPC and one of 2 SNPs associated with 1-(1-enyl-stearoyl)- 2-arachidonoyl-GPE, is in near perfect linkage disequilibrium (r^2^=0.98 in Europeans) with rs174547 and rs174546. These SNPs’ minor T alleles have been shown to increase the risk of aortic stenosis European (25) and coronary artery disease and ischemic stroke in Chinese populations (26), respectively. Such an increase is consistent with our results, where our genetic and ASCVD associations suggest plasmalogen levels mediate the deleterious effect of FADS1/2 polymorphisms on ASCVD risk.

**Figure 2:**
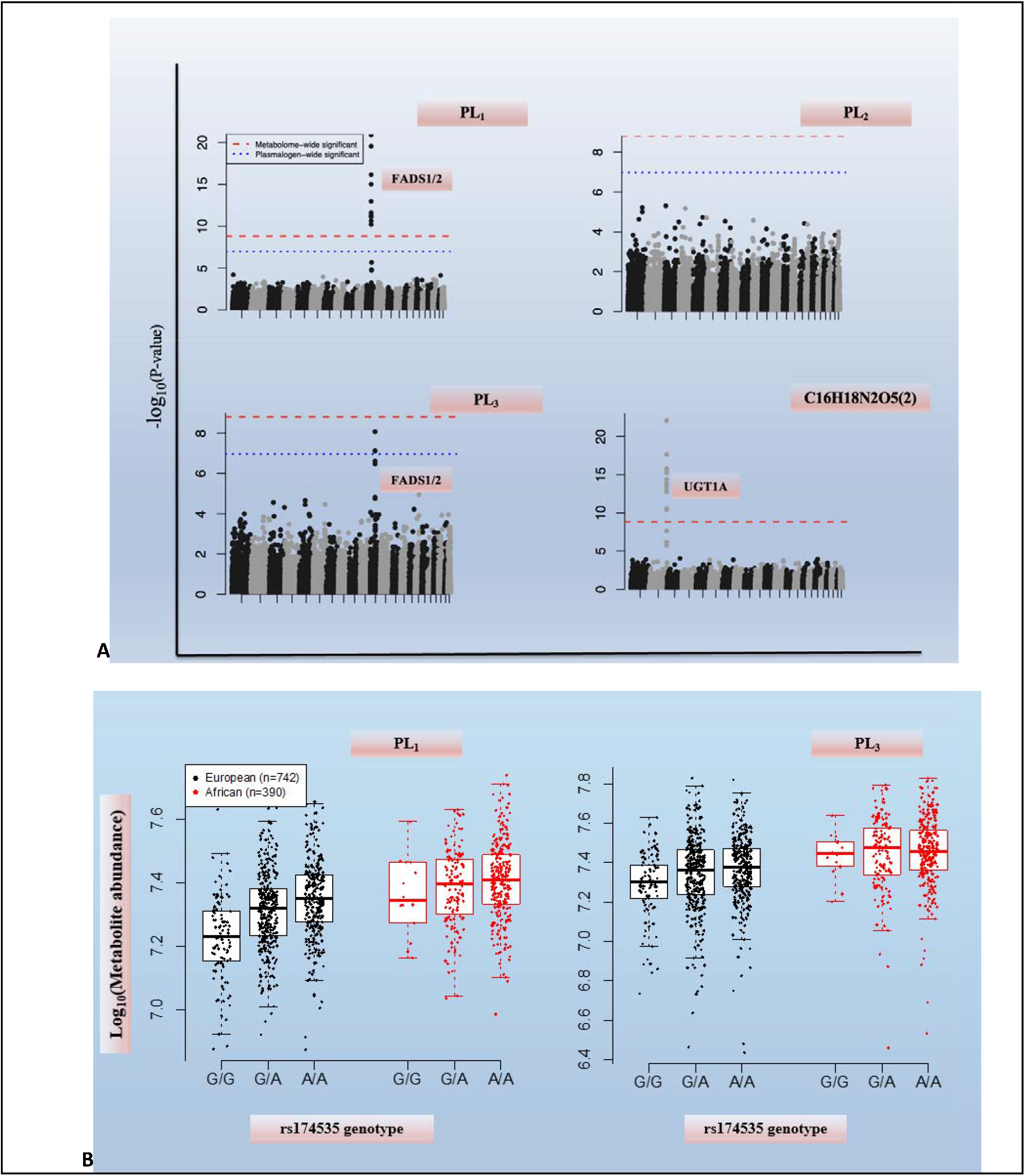
Genetic Influence on Significant Metabolite Species (A) with racial distribution of PL1 and PL2 (B). Abbreviations: PL = Plasmalogens, PL1: 1-(1-enyl-palmitoyl)-2-arachidonoyl-GPC (P-16:0/20:4), PL2: 1-(1-enyl-palmitoyl)-2-arachidonoyl-GPE (P-16:0/20:4),PL3: 1-(1-enyl-stearoyl)-2-arachidonoyl-GPE (P-18:0/20:4)

Our finding of an inverse association of alpha-ketobutyrate with ASCVD events is novel and, to our knowledge, has not been previously reported. Alpha-ketobutyrate is involved in the metabolism of several amino acids, propanoate, and C-5 branched dibasic acid, and is a degradation product of threonine. Walter et al. previously reported that increased hepatic glutathione stress can lead to increased supply of alpha-ketobutyrate. The conversion of alpha-ketobutyrate to its byproduct alpha-hydroxybutyrate indicates early signs of insulin resistance (44). Using proteomics and network analysis to assess vein graft kinetics in mice, Decano et al., showed that tricarboxylic acid substrate utilization of α-ketobutyrate showed a trend of increasing rate on pemafibrate treatment and reversal on PPARα (peroxisome proliferator-activated receptors) silencing resulting in less vein graft lesions and failure (45). It is thus plausible that alpha-ketobutyrate levels form an equilibrium between oxidative stress response and endothelial injury.

We also identified a strong inverse ASCVD association with C_16_H_18_N_2_O_5_, a bilirubin degradation product. This finding is consistent with a UK Biobank study, which reported negative causal associations of total bilirubin levels with cardiovascular disease, and coronary heart disease and the Veterans Aging Cohort Study CVD (46,47). Furthermore, it has been previously reported that chronic hyperbilirubinemia due to Gilbert Syndrome is associated with a substantially lower risk of ischemic heart disease and CVD events (48). These findings may be attributed to bilirubin’s anti-oxidant properties as a superoxide scavenger, modulation of PPAR, and cytoprotective properties (49,50)

Further, we found that the Glucuronosyltransferase Family 1 Member A Complex Locus (UGT1A) gene that regulate bilirubin concentrations had an association with ASCVD outcomes in the present study, which is also consistent with prior literature (46,51-53). Prior genetic studies of bilirubin-associated variants, including those in the UGT1A1 gene, have shown relevance of this molecule to diseased cardiometabolic states. In their experimental work, Takei et al., showed that biliverdin treatment in mice reversed the higher gene expression of CD11c, encoding M1 macrophage marker, and TNF α, encoding the proinflammatory cytokine tumor necrosis factor-α in mice (54). Thus, bilirubin and its metabolites may have a role in reducing active inflammation in adipose tissues (54) and activation of nuclear receptors responsible for burning fats (55) which may directly impact cardiometabolic health.

### Study Limitations

Our study has some notable limitations. First, although we found significant biologically plausible associations between several metabolites and incident ASCVD events that are supported by a genetic association study, our results must be replicated across other cohorts. Second, frozen plasma samples were used for analysis, which could have influenced measured concentrations of metabolites and lipids. However, we used contemporary quantitative lipidomic and metabolomic techniques that are unlikely to be affected by storage (56). Third, more precise contributions of dietary intake, lifestyle, stress, and environmental exposure that may influence metabolomic profiles were not included in this analysis. Finally, we measured metabolites at one time point. Future studies that perform serial metabolomic analysis may provide additional insight into our findings.

## CONCLUSION

Our data indicate that higher mid-life levels of three Plasmalogens, two amino acid metabolites, and a bilirubin degradation product, all of which have anti-inflammatory properties, are associated with lower risk of late-life ASCVD events. The biological plausibility of these observations is supported by our findings that several of these metabolites are directly influenced by genetic polymorphisms, which have previously been associated with ASCVD events. When replicated by other cohorts and mechanistic studies, our findings may have significant implications on ASCVD risk stratification as well as primordial, primary, and secondary ASCVD prevention.

## Data Availability

All data produced in the present study are available upon reasonable request to the authors

## Acknowledgments

The authors acknowledge the contributions of Amy Beto, Janet Bonk, Mary Catherine Coast, Jowanda Green, Carol Hrtyanski, Louise Martin, Lee Ann McDowell, Jennifer Rush, and Roberta Spanos to implementation of the Heart SCORE study.

## Notes

### Competing Interest Statement

The authors have declared no competing interest.

### Funding Statement

This study was funded by the Pennsylvania Department of Health (ME-02-384). The department specifically disclaims responsibility for any analyses, interpretations, or conclusions. Additional funding was provided by National Institutes of Health grant R01HL089292.

### Author Declarations

IRB Approval need was waived.

