## Supplemental Figures and Table for "Mid-Life Plasmalogens and Other Metabolites with Anti-Inflammatory Properties are Inversely Associated with Long term Cardiovascular Disease Events: Heart SCORE Study"

**Supplemental Tables and Figures**

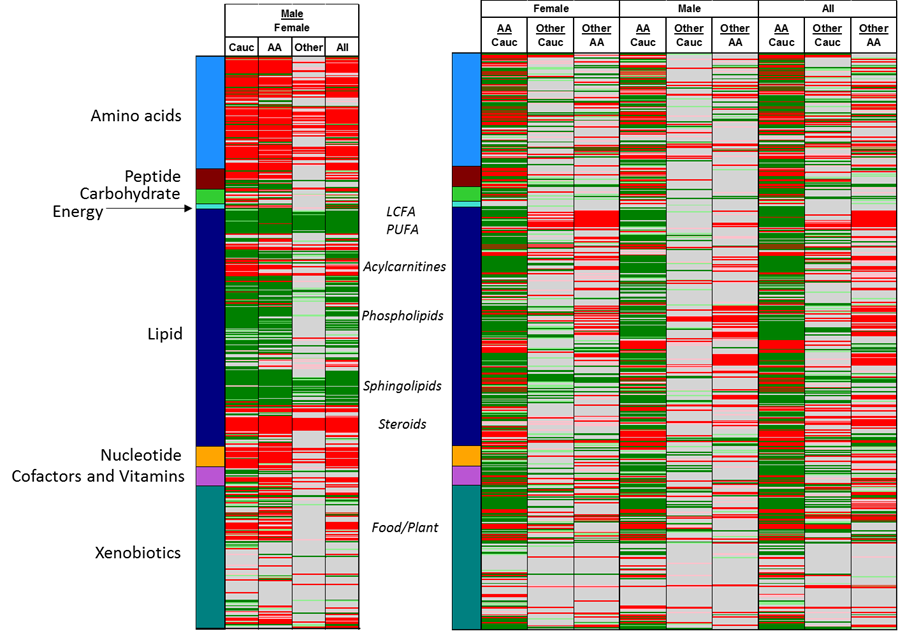
**Figure i: Condensed heat maps showing wide scale differences in Gender and Race groups**

**Table i: Means of significant metabolites distribution and their subclasses in Black and White participants**

| **Significant Metabolite** | **Sub_Pathway** | **Black** | **White** | **p.value** |
| --- | --- | --- | --- | --- |
| X - 16946 | NA | 8.142936512 | 8.250412934 | 8.99E-09 |
| 1-(1-enyl-palmitoyl)-2-arachidonoyl-GPC (P-16:0/20:4)* | Plasmalogen | 10.6752593 | 10.55616375 | 1.63E-42 |
| alpha-ketobutyrate | Methionine, Cysteine, SAM and Taurine Metabolism | 9.219190614 | 9.268761129 | 7.08E-05 |
| pentadecanoate (15:0) | Long Chain Fatty Acid | 11.05609448 | 11.13381177 | 1.66E-12 |
| X - 24295 | NA | 8.837034865 | 8.876304746 | 0.007947478 |
| 1-(1-enyl-palmitoyl)-2-arachidonoyl-GPE (P-16:0/20:4)* | Plasmalogen | 10.08953639 | 9.944264834 | 3.27E-29 |
| 1-(1-enyl-stearoyl)-2-arachidonoyl-GPE (P-18:0/20:4)* | Plasmalogen | 10.74575594 | 10.59320508 | 1.95E-33 |
| 1-palmitoyl-2-linoleoyl-GPI (16:0/18:2) | Phosphatidylinositol (PI) | 8.278687967 | 8.314023086 | 0.014032455 |
| 1-stearoyl-2-oleoyl-GPE (18:0/18:1) | Phosphatidylethanolamine (PE) | 9.174636035 | 9.21692591 | 0.022524244 |
| 2-oxoarginine* | Urea cycle; Arginine and Proline Metabolism | 7.804232882 | 7.779274544 | 0.040768935 |
| X - 14056 | NA | 10.00447548 | 9.942293885 | 6.96E-06 |
| X - 19438 | NA | 8.232015581 | 8.251886783 | 0.096568826 |
